# Survival among cancer patients after a coalmine fire: Analysis of registry data in regional Victoria, Australia

**DOI:** 10.1101/2024.05.19.24307600

**Authors:** Tyler J Lane, Pei Yu, Caroline Gao, Catherine L Smith, Sherene Loi, Natasha Kinsman, Jill Ikin, Yuming Guo, Malcolm R. Sim, Michael J. Abramson

**Affiliations:** School of Public Health and Preventive Medicine, Monash University, Melbourne VIC Australia; Orygen, Centre for Youth Mental Health, The University of Melbourne, Parkville VIC Australia; Peter MacCallum Cancer Centre, Melbourne VIC Australia

## Abstract

**Background:** Air pollution is associated with poorer survival among cancer patients. However, much of the evidence is from studies of ambient air pollution rather than acute exposures like from fires. In this study, we examined the effect of a 2014 coalmine fire in regional Australia, which shrouded nearby communities in smoke for six weeks.

**Methods:** We analysed Victorian Cancer Registry data on new cancers diagnosed between January 2009 and February 2014, the start of the coalmine fire, and survival up to August 2019. Tumours were grouped by location and subtypes for breast and lung cancers. The exposure group was based on residence at diagnosis: Morwell (most exposure), the rest of Latrobe Valley (less exposure), and the rest of Victoria excluding Melbourne (little to no exposure). Those who were diagnosed in Morwell or the Latrobe Valley but died before the fire were classified as unexposed. Months of survival post-fire were compared using Cox proportional hazards frailty and discrete survival models.

**Results:** In Morwell 488 total cancers were diagnosed, 1738 in the rest of the Latrobe Valley, while 42,738 were unexposed. There was no detectable overall effect. However, there were significant reductions in survival among cancers of female reproductive organs in Morwell and breast in the rest of Latrobe Valley.

**Conclusion:** There were no overall changes in cancer survival and isolated increases in two subtypes, although not consistent across exposure sites. Reduced survival from female reproductive cancers were plausibly related to smoke exposure, though numbers were small and there is a high likelihood this was a chance finding. Otherwise, we found little evidence that medium-duration exposure to smoke from the Hazelwood coalmine fire shortened survival among cancer patients. However, owing to limited statistical power, we could not rule out an effect of smoke on cancer survival.

## 1 Introduction

Air pollution shortens the lives of people living with cancer.^1^ The effect is most consistently detected in lung cancers^1,2^ although recent systematic reviews suggest increased mortality in liver, pancreatic, laryngeal, colorectal, bladder and kidney cancers.^1,3^ Some subtypes based on morphology or receptor status may be more responsive to air pollution. For instance, the largest detectable effects of air pollution on lung cancers appear to be among adenocarcinomas.^4,5^

There are several proposed mechanisms to explain how air pollution affects cancer survival. The first is by directly acting on existing tumours. Fine particulate matter (e.g., PM_2.5_) and gaseous air pollution (e.g., volatile organic compounds, aldehydes, nitrogen oxides, polycyclic aromatic hydrocarbons and metals) can activate the nuclear factor erythroid 2 (NRF2) gene, promoting cancer cell growth and resistance to chemotherapy.^6^ Another mechanism is, rather than acting on existing tumours, the air pollution causing tumours with poorer prognoses, such as breast cancers that are triple negative for receptors (HER2, ER, PR).^7,8^ The effects of air pollution on cancer survival may also be indirect, for instance by increasing the risk of cardiopulmonary death in people whose health has been compromised by cancer.^9^

Air pollution may be more harmful when originating from large-scale combustion events like wildfires. Even when using standard measurements like particulate matter <2.5µm (PM_2.5_), wildfire smoke contains greater amounts of smaller particulate matter that can penetrate deeper into the lungs and are more toxic due to higher concentrations of oxidative and pro-inflammatory components.^10,11^ Coal fires may have additional risk compared to other fires as coal combustion releases components associated with poorer cancer survival including heavy metals like cadmium and arsenic and gases such as NO_2_ and O_3_.^4,12–15^ These components may also increase the likelihood of cardiopulmonary events that potentially lead to earlier mortality in cancer patients.^2,10^

In this study, we examined whether cancer survival was affected by the 2014 Hazelwood coalmine fire in regional Victoria, Australia, which shrouded the nearby town of Morwell and the wider Latrobe Valley in smoke and ash for six weeks.^16^

In this study, we addressed the following questions:

1. Did air pollutant exposure from the Hazelwood coal mine fire shorten survival among people with existing cancer diagnoses?
2. Did any effects identified vary by cancer site or subtype?

## 2 Methods

### 2.1 Cancer data

This study analysed Victorian Cancer Registry data. Since 1982, all public and private hospitals, pathology services, public health services, and radiation therapy centres in the State of Victoria have been required to report all newly-diagnosed cancers to the registry.^17^ Registry data are linked to the Victorian Registry of Births Deaths and Marriages as well as the National Death Index. Tumours are classified based on topography and morphology using the International Classification of Diseases for Oncology (ICD-O). More information about the Victorian Cancer Registry can be found in their annual report.^18^

Breast cancer subtypes based on receptor status including human epidermal growth factor positive (HER2+), oestrogen receptor and/or progesterone receptor positive (combined into ER+/PR+), and triple negative for receptors, were derived from data included in the registry. Only those with a negative test for all three receptors were classified triple negative. Lung cancer subtypes included small and large cell carcinoma, adenocarcinoma, and squamous cell carcinoma, based on International Agency for Research on Cancer categorisation of ICD-O morphology codes.^19,20^ Specific codes for cancer locations and morphologies are available in Table S1 in the Supplementary materials.

### 2.2 Inclusion/exclusion criteria

Records were limited to invasive cancers with at least one day of survival, diagnosed between January 2009 and February 2014 prior to the start of the coal mine fire. Cancers diagnosed after the fire were excluded since they could not be reliably classed as smoke-exposed. Only records with valid residential information that could be linked to geographic data were included. Data were right-censored after August 2019 to account for potential confounding from the 2019/2020 “Black Summer”, an unprecedented bushfire season in terms of intensity and duration that covered much of south-eastern Australia in smoke.^21^

We excluded the Melbourne metropolitan area due to substantial sociodemographic and health differences that we did not believe could be adequately adjusted for in regression analyses. These differences included a 10% higher age-adjusted cancer mortality rate in regional Victoria compared to Melbourne.^18^

### 2.3 Exposure

Due to insufficient air monitoring data available during the mine fire period, exposure to the fire-related PM_2.5_ concentrations was retrospectively modelled using meteorological, dispersion, and chemical transport modelling.^22^ As illustrated in Figure 1, the coalmine is located in the Latrobe Valley local government area (see Figure S1 in the Supplementary Materials for illustrations of fire-related PM_2.5_ distributions across Victoria). Estimated exposure was highest in Morwell (population 13,771 as of the 2016 Census), which was adjacent to the coalmine.

**Figure 1.**
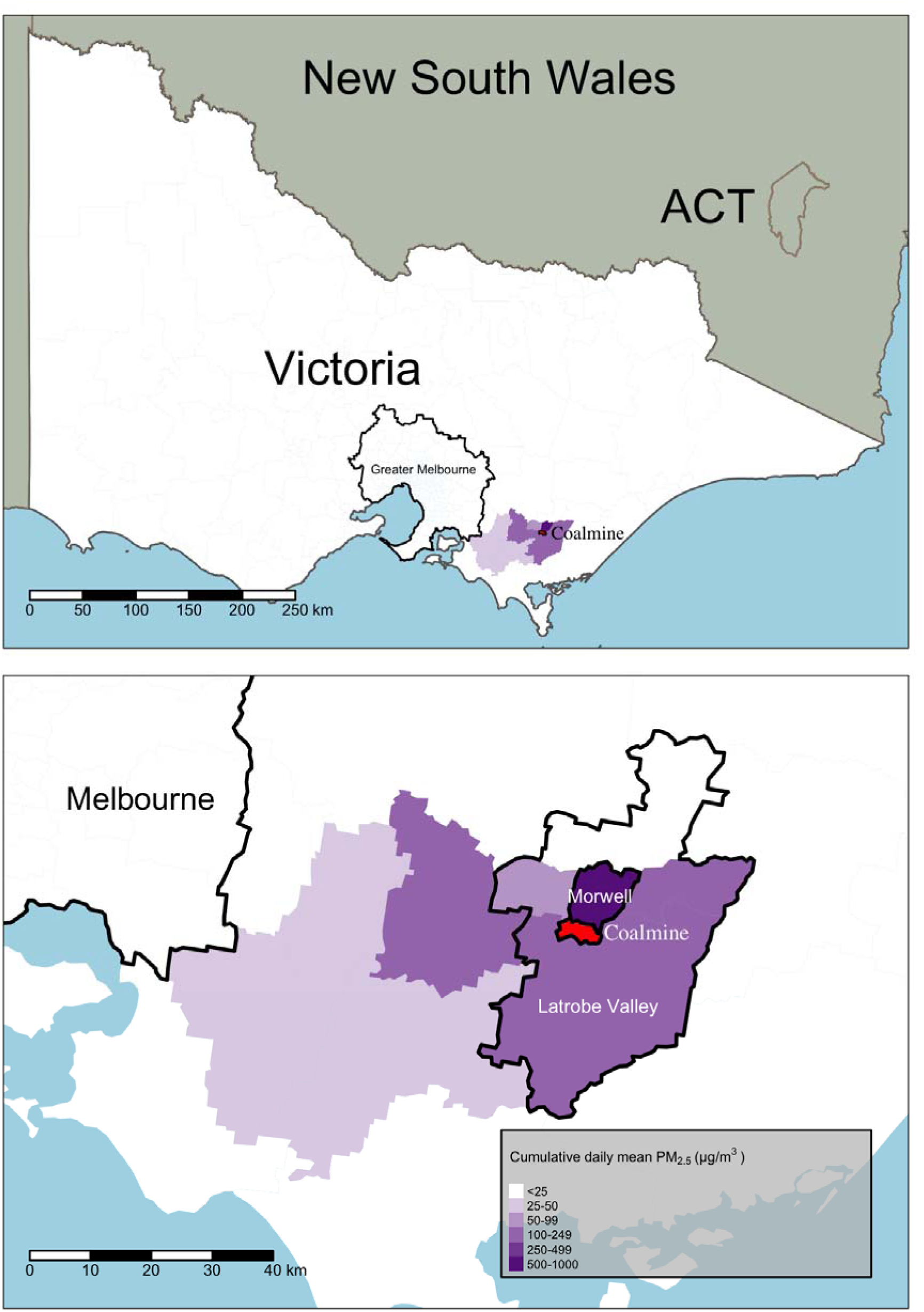
Map of Australia showing the location of the coalmine and of the surrounding areas, with modelled PM_2.5_ distributions from the 2014 coalmine fire; see Figure S1 for a full map of fire-related PM_2.5_ throughout Victoria

The Environment Protection Authority Victoria classifies air quality as “good” when daily mean particulate matter <2.5 µm (PM) has a concentration below 12.5µg/m^3^ and “extremely poor” when it exceeds 150µg/m^3^.^23^ Air quality models estimated that daily mean PM reached 1022µg/m^3^ in residential areas of Morwell that were near the mine during the fire.^22^ Across Morwell, the population-weighted daily mean of fire-related PM was 19.0µg/m^3^ over the entire course of the fire (9 February to 25 March 2014) and 32.8µg/m^3^ over the peak period (9 February to 6 March 2014; see Figure S2 for daily PM_2.5_ estimates). Other areas of the Latrobe Valley (population 59,328 as of the 2016 Census, excluding Morwell^24^) received smoke as well, with population-weighted daily mean of 1.8µg/m^3^ for the entire mine fire period and 3.1µg/m^3^ during the peak.

Residents of other Latrobe Valley towns also have similar socio-demographic characteristics and similar exposures to other air pollutants (e.g., Hazelwood open-cut mine and power station) compared with Morwell, although aside from Moe/Newborough, Morwell is considerably more socioeconomically disadvantaged.^25^ The rest of the Latrobe Valley is considerably larger and provides better statistical power, though smoke exposure was diluted. Therefore, if the fire affected cancer survival, we expected larger but less precise estimates in Morwell and more precise but smaller estimates in the rest of the Latrobe Valley.

Therefore, we categorised exposure time into three groups: (1) highly exposed – time post-fire among cancer patients living in Morwell; (2) moderately exposed – time post-fire among cancer patients living in the rest of Latrobe Valley; and (3) little to no exposure – all survival time among cancer patients living in other regional areas in Victoria, and survival time prior to the mine fire among cancer patients living in Morwell or rest of Latrobe Valley.

### 2.4 Statistical analysis

To ensure the robustness of results and control for spatial clustering, we used two types of mixed-effects survival models: Cox proportional hazards frailty^26^ and mixed-effects discrete-time survival.^27^ In both models, survival was evaluated monthly; we used a logit link to fit the discrete-time survival model. Random effects were included for each Statistical Area at level 2 (SA2, which approximate town boundaries in regional areas) to account for area-related variations in cancer survival that preceded the mine fire and were due to other factors that are not controlled in the model such as access to services, demographics, and lifestyle. The Hazelwood coalmine fire was treated as a time-varying exposure (as described in section 2.3). Analyses were adjusted for the following confounders: age at diagnosis (converted to a natural spline with five degrees of freedom to account for non-linearities), sex, tumour grade (high, other), year of diagnosis, and area-based indicator of socioeconomic status measured using Index of Relative Socio-economic Advantage and Disadvantage (IRSAD).^25^ Analyses were conducted in R^28^ using RStudio.^29^ Cox proportional hazards frailty models were fitted with the *coxph* function in the *survival* package^30^ and mixed effects discrete survival models were fitted using the *bam* function in the *mgcv* package^31^ with random intercepts estimated using the smooth function.

### 2.5 Ethics approval

This study was approved by the Monash University Human Research Ethics Committee (MUHREC) as part of the Hazelwood Adult Survey & Health Record Linkage Study on 21 May 2015 (CF15/872 – 2015000389). The approval was extended on 7 September 2020 (25680). Access to the Victorian Cancer Registry was approved by the Director on 6 May 2016 (CF15/187).

## 3 Results

### 3.1 Descriptives

From January 2009 to February 2014, 44,964 new cancers were diagnosed in regional Victoria, where the patient lived at least one day after diagnosis (2139 had no days after diagnosis). Table 1 presents counts for each cancer type by exposure site (Morwell, rest of Latrobe Valley, and rest of regional Victoria).

**Table 1.** Cancer cases from January 2009 to February 2014 by exposure site, location and, for breast cancers, receptor subtype.

| Cancer location and type | Morwell | Rest of Latrobe Valley | Rest of regional Victoria |
| --- | --- | --- | --- |
|  | N (rate per 10,000 person years)* |  |  |
| All cancers | 488 (6.65) | 1738 (5.69) | 42,738 (6.37) |
| Blood (lymphoid, haematopoietic, and related issues) | 53 (0.72) | 210 (0.69) | 4673 (0.70) |
| Brain & other neurological | 3 (0.04) | 28 (0.09) | 655 (0.10) |
| Breast | 60 (0.82) | 195 (0.64) | 5096 (0.76) |
| HER2+ | 6 (0.08) | 34 (0.11) | 752 (0.11) |
| ER+/PR+ | 46 (0.63) | 149 (0.49) | 4075 (0.61) |
| Triple negative | 8 (0.11) | 24 (0.08) | 469 (0.07) |
| Undetermined/untested | 0 (0) | 8 (0.03) | 176 (0.03) |
| Digestive organs | 115 (1.57) | 350 (1.15) | 9224 (1.37) |
| Female reproductive organs | 27 (0.37) | 80 (0.26) | 1834 (0.27) |
| Male reproductive organs | 65 (0.89) | 283 (0.93) | 7154 (1.07) |
| Melanoma | 28 (0.38) | 141 (0.46) | 3840 (0.57) |
| Mesothelioma | 7 (0.10) | 20 (0.07) | 217 (0.03) |
| Oral (lip, oral cavity, pharynx) | 18 (0.25) | 57 (0.19) | 1261 (0.19) |
| Respiratory and intrathoracic organs | 62 (0.84) | 200 (0.66) | 4228 (0.63) |
| Lung | 56 | 186 | 3905 |
| Adenocarcinoma | 15 (0.20) | 60 (0.20) | 1366 (0.2) |
| Large cell carcinoma | 8 (0.11) | 13 (0.04) | 301 (0.04) |
| Small cell carcinoma | 4 (0.05) | 16 (0.05) | 441 (0.07) |
| Squamous cell carcinoma | 10 (0.14) | 44 (0.14) | 772 (0.12) |
| Adenosquamous† | 0 (0) | 0 (0) | 28 (0.00) |
| Undetermined | 19 (0.26) | 53 (0.17) | 997 (0.15) |
| Thyroid & other endocrine glands | 5 (0.07) | 20 (0.07) | 550 (0.08) |
| Urinary tract | 28 (0.38) | 94 (0.31) | 2248 (0.34) |
| All others (others and unknown) | 17 (0.23) | 60 (0.20) | 1758 (0.26) |

\* Person years calculated for each area by summing the estimated annual populations from the Australian Bureau of Statistics for the years 2009 to 2014 (population figures for 2014 were multiplied by 2/12<sup>ths</sup> to account for inclusion of cancers only if they were diagnosed in the first two months of the year); these were 73,438 for Morwell, 305,290 for the rest of Latrobe Valley, and 6,709,443 for the rest of regional Victoria
† Adenosquamous cancers were included in analyses of both adenocarcinoma and squamous cell carcinomas rather than analysed separately

### 3.2 Cancer survival after the Hazelwood coalmine fire

#### 3.2.1 Overall and by major site

Effects were largely consistent between modelling approaches in terms of magnitude, direction, and precision of estimates (Figure 2). There was no detectable effect of the mine fire on cancer survival overall and significant associations with only two cancer types. The first statistically significant association was shorter survival among patients with female reproductive organ cancers in Morwell (Cox PH – RR: 3.50 [95% CI: 1.11-11.10, *p* = 0.033]; discrete survival – RR: 3.28 [95% CI: 1.03-10.49, *p* = 0.045]); there was no evidence of a corresponding pattern in the rest of the Latrobe Valley. The second effect was shorter survival among breast cancers diagnosed in the rest of the Latrobe Valley (Cox PH – RR: 1.94 [95% CI: 1.00-3.76, *p* = 0.049]; discrete survival – RR: 1.89 [95% CI: 0.97-3.68, *p* = 0.060]). However, while point estimates and confidence interval precisions were similar across the models, the association was only significant in the Cox PH frailty model; there also was no corresponding pattern in Morwell. These results are summarised in Figure 2 and Table S2 in the supplementary materials.

**Figure 2.**
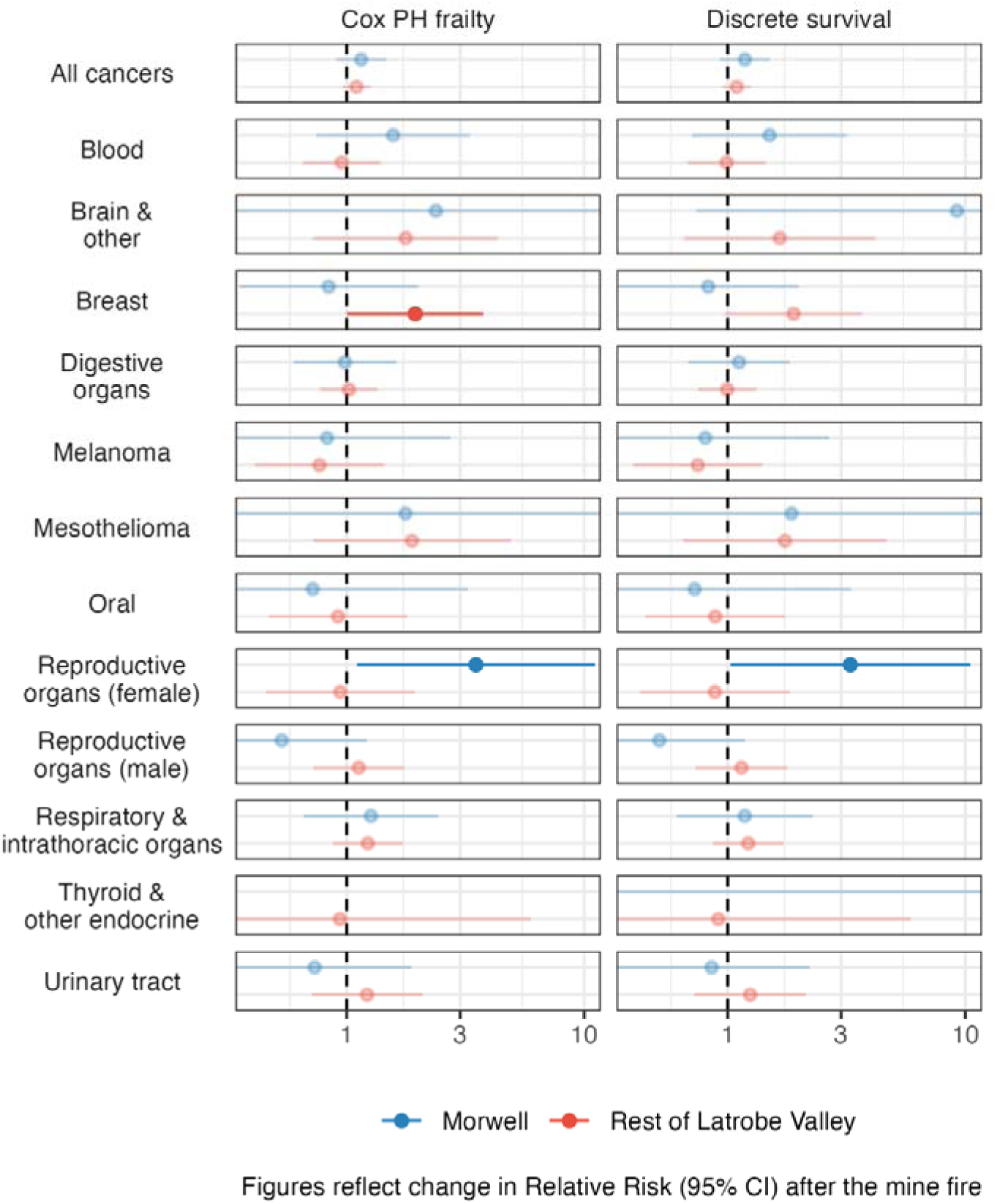
Changes in survival among cancer patients after the Hazelwood coalmine fire; thyroid & other endocrine cancer results for Morwell not shown due to small numbers

#### 3.2.2 Breast and lung cancer subtypes

While survival was shortened among breast cancers overall in the rest of the Latrobe Valley following the mine fire, there were no detectable effects by receptor subtypes (Figure 3). In the rest of Latrobe Valley, point estimates were elevated for triple negative and, to a lesser extent, ER+/PR+ breast cancers, though these were not statistically significant and there was no corresponding pattern in Morwell.

**Figure 3.**
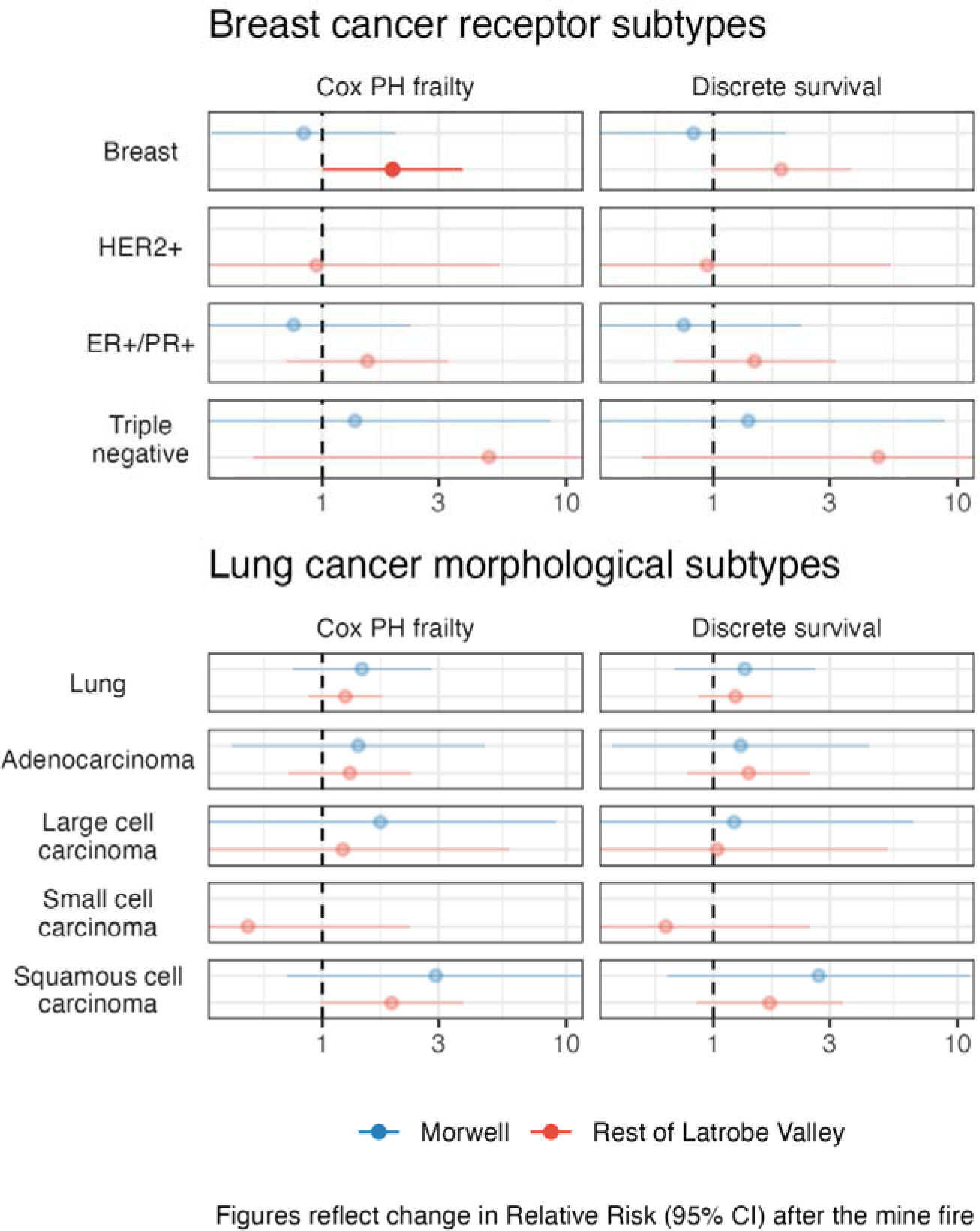
Changes in survival among breast and lung cancer patients after the Hazelwood coalmine fire; HER2+ breast cancers and small cell lung carcinomas in Morwell not shown due to small numbers

There were no detectable effects in lung cancer overall or its morphological subtypes. However, point estimates were noticeably elevated for squamous cell carcinoma, and followed the pattern we would have expected if smoke from the coalmine fire shortened cancer survival: a larger but less precise effect was observed in Morwell. Numbers of both HER2+ breast cancers and small cell lung carcinomas were too small in Morwell to conduct meaningful analyses. These results are summarised in Figure 3 and Table S1 in the supplementary materials.

## 4 Discussion

We found no evidence that the Hazelwood coalmine fire affected overall cancer survival. There was also little evidence of effects on cancer subtypes, with the exception of worse survival among: 1) breast cancers in the rest of Latrobe Valley but not Morwell; and 2) female reproductive organ cancers in Morwell, but not the rest of Latrobe Valley. The first association is likely a chance finding for two reasons: the effect was only detectable in the lower-exposure group (Latrobe Valley), which itself was only marginally significant in one of the two modelling approaches, while the point estimate in the higher-exposure group (Morwell) was in the opposite direction.

The second association between mine fire exposure and shortened female reproductive organ cancer survival could more plausibly be causal since the effect was only detectable in the highest exposure group, Morwell residents. There is also an evidence base linking air pollution to poorer survival in both cervical^32^ and ovarian cancers.^33^ However, this evidence focuses on ambient air pollution which, as a chronic exposure, makes it difficult to disentangle whether the effect was due to inducing tumours with poorer prognoses or worsening outcomes in existing tumours. Further, there was no evidence of a similar effect in the rest of Latrobe Valley. Either the level of exposure in the rest of the Latrobe Valley was too small to have a meaningful or detectable effect on survival among cancers of female reproductive organs, or the effect in Morwell was due to chance (Type 1 error), which seems likely given the marginal statistical significance of this association. It is also more likely than not that we produced at least Type 1 error, with a likelihood of 49% with an alpha of 0.05 and 13 main outcomes.

While there was no detectable change in survival among lung cancer morphological subtypes, point estimates among squamous cell carcinomas followed the expected pattern: larger but less precise in Morwell than the rest of the Latrobe Valley. However, the numbers were small and, with numerous analyses, the likelihood that one group would follow this pattern by chance is high. Further, this association did not correspond to other studies, which reported larger effects among adenocarcinomas.^4,5^

Our findings do not rule out the harms of coalmine fire on cancer survival. This is most notable for respiratory/intrathoracic cancers, which affect organs that have direct exposure to air pollution and where there is a body of evidence indicating a negative effect on survival.^1,2^ The length of exposure, while medium duration, may not have been long enough to affect survival in any detectable way. However, the effects of smoke exposure from fire events likely remain a major risk for cancer patients by virtue of their wide exposure. In the last decade, billions were exposed to smoke from landscape fires,^34^ and probably billions more in this decade due to widespread extreme smoke events like the 2019-2020 Australian “Black Summer” and 2023 Canadian wildfires. There will be no shortage of opportunities to study the effects of smoke on cancer survival with more statistical power, especially as climate change increases the frequency, intensity, and duration of fire events.

### 4.1 Strengths and limitations

This study has several strengths. The use of population-level data for all cancers in regional Victoria, Australia allowed us to include all diagnosed cancers. And, two different modelling approaches, found similar results in terms of magnitude, direction, and significance. The incorporation of random effects by residential area accounted for some residual confounding between sites.

However, there were also several limitations. Statistical power was limited due to low counts, particularly for rare cancers like blood, brain and other neurological, breast subtypes, melanoma, respiratory and intrathoracic organs, and thyroid and other endocrine gland cancers. We lacked individual-level data on socioeconomic status, a potentially major confounder. Exposure was misclassified in some cases since it was based on residence at diagnosis and did not account for relocations. We used a categorical exposure indicator because we lacked more accurate location information during the mine fire that could be matched to granular air pollution estimates. We also lacked data on important lifestyle confounders such as alcohol drinking and smoking tobacco.

## 5 Conclusion

We found minimal evidence that the Hazelwood coalmine fire affected cancer survival. While there were a few significant associations within some cancer subtypes, each was characterised by marginal significance among multiple analyses and could be due to chance. Since these analyses had limited statistical power due to low event counts, they do not preclude a possible effect of extreme smoke exposure on cancer survival for some cancer sub types. As fire events become more frequent and intense due to climate change, there are potentially increasing landscape fire-associated cancer risks at population level, which requires studies at larger scales to address this question with greater statistical power.

## Data Availability

All data are confidential and cannot be shared.

