## Supplementary materials for "Survival among cancer patients after a coalmine fire: Analysis of registry data in regional Victoria, Australia"

### Fire-related smoke distributions

In this section, we build on the map in Figure 1, which focuses on the location of the coalmine fire and the Latrobe Valley as well as smoke distributions, using population-weighted fire-related $PM_{2.5}$ data from Luhar et al. (2020).[22] Figure S1 shows the distributions of cumulative daily mean of population-weighted fire-related $PM_{2.5}$ at Statistical Area Level 2 across Victoria. As can be seen, Morwell (red) is at the extreme end of exposure, as is the Latrobe Valley (yellow), though to a lesser extent.

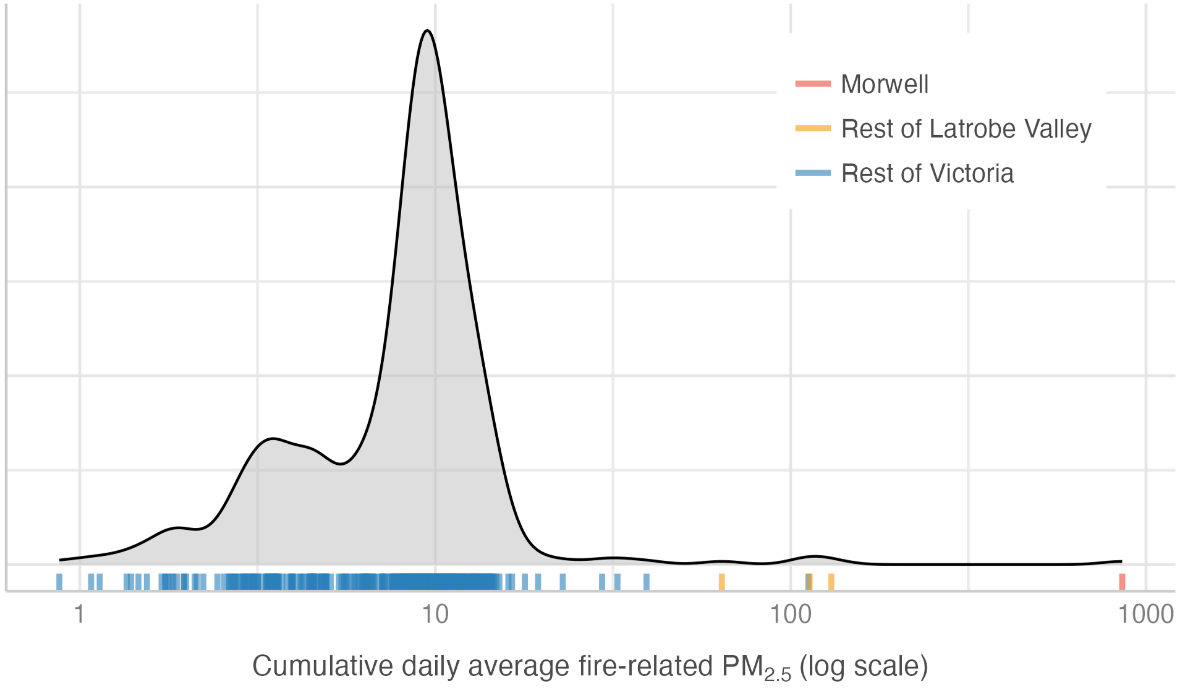

**Figure S2. Distribution of cumulative daily mean $PM_{2.5}$ from the Hazelwood coalmine fire at Statistical Area Level 2, colour-coded by location (Morwell, rest of Latrobe Valley, rest of Victoria); note that the x-axis is logged**

### Smoke distribution over the Hazelwood coalmine fire

Over the course of the Hazelwood coalmine fire, estimated $PM_{2.5}$ distributions were considerably higher in the early phase, particularly in Morwell. This is illustrated in Figure S2. Data are from Luhar et al. (2020).[22]

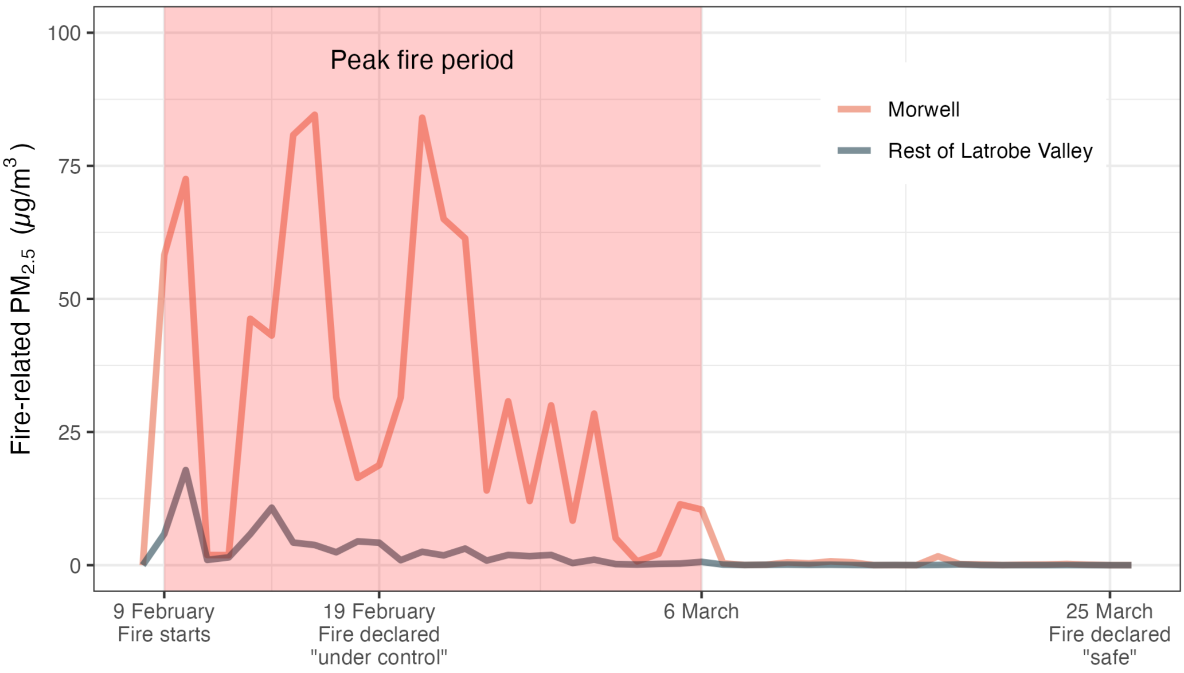

**Figure S2. Modelled daily mean $PM_{2.5}$ from the 2014 Hazelwood coalmine fire in Morwell and the rest of the Latrobe Valley**

### Cancer codes

**Table S1. ICD-10 codes for cancer categories in this paper**

| Cancer Categories (ICD-O codes[35]) |
|---|
| **Lip, oral cavity and pharynyx (C00-C14)** |
| **Digestive organs (C15-C25)** |
| Oesophagus (C15) |
| Stomach (C16) |
| Colorectal (C18-C21) |
| Colon (C18) |
| Rectum (C20) |
| Liver (C22) |
| Pancreas (C25) |
| **Respiratory and intrathoracic organs (C30-C38)** |
| Larynx (C32) |
| Lung (C33-C34) |
| Squamous cell carcinoma* |
| Adenocarcinoma* |
| Large cell carcinoma* |
| Small cell carcinoma* |
| Others* |
| **Melanoma (C43)** |
| **Mesothelioma (C45)** |
| **Breast (C50)** |
| Estrogen Receptor (ER+)† |
| Progesterone Receptor (PR+)† |
| Human Epithelial Growth Factor Receptor (HER2)† |
| Triple negative* |
| **Female reproductive organs (C51-C58)** |
| Cervix (C53) |
| **Male reproductive organs (C60-C63)** |
| Prostate (C61) |
| Testis (C62) |
| **Urinary tract (C64-C68)** |
| Kidney (C64) |
| Bladder (C67) |
| **Brain and other CNS (C70-C72)** |
| Brain (C71) |
| **Thyroid and other endocrine glands (C73-C75)** |
| Thyroid (C73) |
| **Unknown site (C76-C80,C26,C39)** |
| **Lymphoid, haematopoietic + related tissue (C81-C96, D45, D46, D47)** |
| Hodgkins disease (C81) |
| Non-hodgkin lymphoma (C82-C85) |
| Follicular non-Hodgkin's lymphoma (C82) |
| Diffuse non-Hodgkin's lymphoma (C83) |
| Multiple myeloma (C90) |
| Leukaemia (C91-C95) |
| Myelodysplastic* (D46) |
| **All other cancers^ (C40-42, C44, C46-49, C69, C97)** |
| **All malignancy (C00-C97, D45-D47)** |

*** Lung cancer subtypes determined by morphology[19,20]; † Breast cancer subtypes determined by hormone receptor tests**

### Results table

**Table S2. Changes in survival among cancer patients after the Hazelwood coalmine fire among residents of Morwell and the rest of the Latrobe Valley compared to regional Victoria**

| Cancer type | Exposure location | Cox PH frailty – RR (95%CI) | Discrete survival – RR (95%CI) |
|---|---|---|---|
| All cancers | Morwell | 1.15 (0.90- 1.47; p = 0.258) | 1.18 (0.93- 1.51; p = 0.180) |
| | Rest of Latrobe Valley | 1.10 (0.96- 1.26; p = 0.169) | 1.09 (0.95- 1.25; p = 0.205) |
| Blood | Morwell | 1.57 (0.75- 3.30; p = 0.235) | 1.50 (0.71- 3.18; p = 0.289) |
| | Rest of Latrobe Valley | 0.95 (0.65- 1.39; p = 0.807) | 0.99 (0.68- 1.45; p = 0.958) |
| Brain & other | Morwell | 2.37 (0.21- 27.27; p = 0.488) | 9.21 (0.74- 114.96; p = 0.085) |
| | Rest of Latrobe Valley | 1.77 (0.72- 4.35; p = 0.212) | 1.66 (0.66- 4.18; p = 0.286) |
| Breast | Morwell | 0.84 (0.35- 2.00; p = 0.693) | 0.83 (0.35- 1.98; p = 0.671) |
| | Rest of Latrobe Valley | **1.94 (1.00- 3.76; p = 0.049)** | 1.89 (0.97- 3.68; p = 0.060) |
| Breast – HER2+ | Morwell | NA | NA |
| | Rest of Latrobe Valley | 0.95 (0.17- 5.31; p = 0.951) | 0.94 (0.17- 5.31; p = 0.946) |
| Breast – ER+/PR+ | Morwell | 0.76 (0.25- 2.31; p = 0.632) | 0.75 (0.25- 2.29; p = 0.620) |
| | Rest of Latrobe Valley | 1.53 (0.72- 3.28; p = 0.270) | 1.47 (0.69- 3.16; p = 0.320) |
| Breast – Triple negative | Morwell | 1.36 (0.22- 8.65; p = 0.742) | 1.39 (0.22- 8.86; p = 0.730) |
| | Rest of Latrobe Valley | 4.83 (0.52- 44.72; p = 0.166) | 4.74 (0.51- 44.16; p = 0.172) |
| Digestive organs | Morwell | 0.98 (0.60- 1.62; p = 0.946) | 1.12 (0.68- 1.82; p = 0.661) |
| | Rest of Latrobe Valley | 1.02 (0.77- 1.35; p = 0.878) | 1.00 (0.75- 1.32; p = 0.972) |
| Melanoma | Morwell | 0.83 (0.25- 2.73; p = 0.755) | 0.81 (0.24- 2.68; p = 0.724) |
| | Rest of Latrobe Valley | 0.77 (0.41- 1.43; p = 0.407) | 0.75 (0.40- 1.40; p = 0.366) |
| Mesothelioma | Morwell | 1.77 (0.19- 16.48; p = 0.617) | 1.86 (0.19- 18.03; p = 0.594) |
| | Rest of Latrobe Valley | 1.88 (0.72- 4.90; p = 0.196) | 1.74 (0.65- 4.66; p = 0.271) |
| Oral | Morwell | 0.72 (0.16- 3.25; p = 0.669) | 0.73 (0.16- 3.30; p = 0.679) |
| | Rest of Latrobe Valley | 0.92 (0.47- 1.80; p = 0.806) | 0.89 (0.45- 1.74; p = 0.728) |
| Reproductive organs (female) | Morwell | **3.50 (1.11- 11.10; p = 0.033)** | **3.28 (1.03- 10.49; p = 0.045)** |
| | Rest of Latrobe Valley | 0.94 (0.46- 1.93; p = 0.867) | 0.88 (0.43- 1.83; p = 0.738) |
| Reproductive organs (male) | Morwell | 0.53 (0.23- 1.21; p = 0.134) | 0.52 (0.23- 1.18; p = 0.117) |
| | Rest of Latrobe Valley | 1.12 (0.72- 1.74; p = 0.603) | 1.14 (0.73- 1.78; p = 0.556) |
| Respiratory & intrathoracic organs | Morwell | 1.26 (0.66- 2.42; p = 0.479) | 1.18 (0.61- 2.28; p = 0.624) |
| | Rest of Latrobe Valley | 1.23 (0.87- 1.72; p = 0.238) | 1.22 (0.87- 1.72; p = 0.255) |
| Lung | Morwell | 1.45 (0.76- 2.80; p = 0.263) | 1.34 (0.69- 2.61; p = 0.386) |
| | Rest of Latrobe Valley | 1.24 (0.88- 1.76; p = 0.219) | 1.23 (0.87- 1.75; p = 0.246) |
| Lung - *adenocarcinoma* | Morwell | 1.41 (0.43- 4.65; p = 0.577) | 1.29 (0.38- 4.34; p = 0.678) |
| | Rest of Latrobe Valley | 1.30 (0.73- 2.32; p = 0.377) | 1.39 (0.78- 2.50; p = 0.264) |
| Lung – *large cell carcinoma* | Morwell | 1.73 (0.33- 9.08; p = 0.517) | 1.21 (0.22- 6.59; p = 0.822) |
| | Rest of Latrobe Valley | 1.21 (0.25- 5.82; p = 0.808) | 1.03 (0.21- 5.18; p = 0.967) |
| Lung – *small cell carcinoma* | Morwell | NA | NA |
| | Rest of Latrobe Valley | 0.50 (0.11- 2.28; p = 0.368) | 0.64 (0.16- 2.49; p = 0.517) |
| Lung – *squamous cell carcinoma* | Morwell | 2.92 (0.72- 11.87; p = 0.135) | 2.70 (0.65- 11.24; p = 0.173) |
| | Rest of Latrobe Valley | 1.93 (0.98- 3.79; p = 0.057) | 1.70 (0.85- 3.38; p = 0.132) |
| Thyroid & other endocrine | Morwell | NA | NA |
| | Rest of Latrobe Valley | 0.94 (0.15- 5.96; p = 0.945) | 0.91 (0.14- 5.88; p = 0.924) |
| Urinary tract | Morwell | 0.73 (0.29- 1.87; p = 0.517) | 0.86 (0.33- 2.21; p = 0.748) |
| | Rest of Latrobe Valley | 1.22 (0.71- 2.09; p = 0.470) | 1.24 (0.72- 2.14; p = 0.431) |
